# The Role of the Clinical Pharmacist in Addressing Drug-Related Problems in Stroke Patients in a Tertiary Care Centre

**DOI:** 10.1101/2024.07.09.24309334

**Authors:** Susan Philip, Jeesa George, A Pramod Kumar, Foujia Begum, K Divya Bharathi

## Abstract

The morbidity, mortality, and diminished quality of life of patients are all impacted by drug-related problems. Stroke is one of the leading causes of disability and death in India and since the stroke patients are at an increased risk of DRPs (Drug Related Problems), early identification, prevention and resolution of the same is important for improving the quality of life of stroke patients. The aim of the study was to assess the incidence of DRPs in stroke patients by using Hepler-strand classification and the acceptance rate by the multidisciplinary team. The relevant information were documented using a predefined data collection form and was analyzed for drug related problems and categorized according to Hepler–Strand classification. A total of 510 DRPs from 130 participants with an average incidence of 3.92 DRPs per patient. Drug-drug interaction was found to be 23.53% and ADRs accounted for 12.55 % of total DRPs. The study found a high acceptance rate of recommendations by healthcare professionals at 97.36%, with changes in therapy being made 68.13% of the time. Early detection and resolution of DRP by clinical pharmacist may improve the therapeutic outcomes in stroke patients. This also helps in preventing complications and unnecessary hospitalization, high cost of treatment and deaths among stroke patients.

## INTRODUCTION

Stroke is one of the leading causes of disability and death in India. World Health Organization (WHO) defined stroke as “Rapidly developing clinical signs of focal (or global) disturbance of cerebral function, lasting more than 24 hours or leading to death, with no apparent cause other than that of vascular origin”^1^. Incidence of stroke increased by 70%, stroke mortality increased by 43%, stroke prevalence increased by 102%, and Disability Adjusted Life Years (DALY) increased by 143% from 1990 to 2019. According to the Global Stroke Factsheet published in 2022, the lifetime risk of having a stroke has climbed by 50% in the last 17 years, with 1 in 4 people being thought to be at risk^2^. Stroke incidence and mortality are higher in Asian countries than in western countries^3^. The disparity between the incidence rates of stroke and coronary heart disease (CHD) in Asian and western populations is usually attributed to higher prevalence of hypertension and a lower level of serum total cholesterol in the Asian population^3,4^. The risk factors for stroke can be classified into nonmodifiable, modifiable, and potentially modifiable. Recommendations for risk factor reduction aggressively target the modifiable risk factors. The most common modifiable, well-documented risk factors for stroke include hypertension, cigarette smoking, diabetes, atrial fibrillation, and dyslipidemia. The nonmodifiable risk factors are age, race, sex, low birth weight, and family history. The potentially modifiable risk factors include oral contraceptives, migraine, drug and alcohol abuse, hemostatic and inflammatory factors and sleep disordered breathing^5^.

In any healthcare setting, ensuring patients’ safety has always been a crucial part of providing care as the complex drug regimen required to treat acute stroke and prevent secondary stroke makes it challenging to prescribe an appropriate drug therapy^6^. Drug related problem (DRP) is defined as an event or circumstance involving drug therapy that actually or potentially interferes with desired health outcomes^7^. Drug related problems are relatively common in hospitalized patients and can result in patient morbidity and mortality, and increased costs. Alongside, the occurrence of a DRP may hinder or postpone the accomplishment of intended therapeutic objectives, worsening patients’ quality of life (QoL)^8^.

Factors associated with drug related problems in stroke patients include advanced age (>65 years), poly pharmacy, comorbid medical conditions, concomitant medications, noncompliance by the patient, lack of proper laboratory and therapeutic drug monitoring, advanced age, altered level of consciousness, impaired communication because of aphasia, invasive nature of diagnostic evaluations, high prevalence of comorbid conditions, co-administration of multiple medications, use of intravenous route of administration because of impaired oral intake, administration of medications that require frequent laboratory testing and dose adjustments, and long hospital stay^9^. The Hepler-Strand classification is one of many that were used to categorize drug-related problems. According to the Hepler-Strand classification, DRPs were categorized into eight groups including untreated indications, subtherapeutic dosage, over dosage, drug use without a clear indication, failure to receive drugs, improper drug selection, drug interactions, and adverse drug reactions^10^.

Clinical pharmacists are skilled in therapeutics and actively involved in putting therapeutic treatment plans into action and keeping an eye on them. They assisted doctors in analyzing pertinent laboratory data, checking medication orders for plausibility, participating in patient rounds, taking drug histories at admission, identifying and resolving administration errors, adverse drug reactions, and drug-drug interactions, as well as encouraging patients to take their prescribed medications as directed^11^.Clinical pharmacist involvement in the stroke care unit has been found to increase identification and tracking of medication errors thereby, reducing the rate of DRP’s. The proportion of clinical pharmacists to patient beds was associated with significant reduction in the mortality rate. Since the stroke patients are at an increased risk of DRP, early identification, prevention and resolution of the same is important for improving the quality of life of stroke patients^12^.

A physician –pharmacist collaborative practice can help to improve the patient health and functioning by providing good patient counselling. By taking into consideration the individual patient’s present conditions, the pharmacist can help them take necessary step towards a better lifestyle and improved medication use. Pharmacists could provide effective patient care through their involvement in pharmaceutical care, leading to improved therapeutic outcomes ^13^.

## AIMS AND OBJECTIVES

The prospective study was conducted to assess the incidence of DRPs in stroke patients by using Hepler-strand classification and the acceptance rate by the multidisciplinary team.

## METHODOLOGY

A prospective study was conducted over a period of 12 months in the Inpatient Department (IPD) of General Medicine and Neurology of a tertiary care centre.

### Method of data collection

A data collection form was designed which contains relevant details such as demographics, past medical history, past medication history, diagnosis, laboratory investigations, therapeutic plan, type of drug related problem (DRP) as per the need of the study. Patients’ treatments were monitored on every day and patients were asked about symptoms when necessary. The relevant data were collected while accompanying the clinician 5 days in a week and also from inpatient medical records and the obtained. The treatment initiated was analyzed for drug related problems. DRPs observed in the patients were categorized according to Hepler–Strand classification and recorded on every day basis. Physicians were provided with the information about DRP’s and also suitable strategies to resolve the DRPs such as drug interactions, adverse drug reactions and dose adjustment. The data was computed using MS Excel and descriptive results were expressed as percentages.

### Study inclusion/exclusion criteria

Patients were included in the study if they met the following criteria: > 18 years of age admitted in the department of medicine and neurology during the study period. Patients visiting the outpatient department of the tertiary care centre, pediatric patients and pregnant women were excluded from the study.

### Statistical Analysis

A sample size of 130 patients were enrolled using Cochran formula (N=Z^2^pq/e^2^) from the retrospective data. A sample size of 74 recommendations to resolve the DRP’s was needed to achieve a power of 80% using an alpha error of 0.05.

## RESULTS

Out of the 130 patients enrolled for the study 90(69.23%) were male and 30 (30.77%) were female. Most number of patients had ischemic stroke (53.85%), followed by hemorrhagic stroke (33.85%).12.3% of patients were diagnosed with Transient ischemic attack(TIA). Majority of patients belonged to the age group 61-70. The youngest patient was in their 20s, diagnosed with ischemic stroke and had heart disease since childhood. More than 70% of patients were older than 55 years of age. Male gender had higher incidence of stroke. Hypertension was the highest risk factor. Other risk factors include diabetes, cigarette smoking, ischemic heart disease (IHD), atrial fibrillation, dyslipidemia, obesity, anemia, alcohol use and other cardiac diseases. (**Table 1**).

**Table 1:** Demographic details of patients.

| Patient characteristics | Number | Percentage (%) |
| --- | --- | --- |
| **Gender** |  |  |
| Male | 90 | 69.23 |
| Female | 30 | 30.77 |
| **Age** |  |  |
| 20-30 | 1 | 0.76 |
| 31-40 | 7 | 5.38 |
| 41-50 | 19 | 14.62 |
| 51-60 | 22 | 16.92 |
| 61-70 | 46 | 35.38 |
| 71-80 | 27 | 20.77 |
| 81-90 | 8 | 6.15 |
| **Type of stroke** |  |  |
| Ischemic stroke | 70 | 53.85 |
| Hemorrhagic stroke | 44 | 33.85 |
| Transient ischemic attack (TIA) | 16 | 12.3 |
| **Risk factors** |  |  |
| Age>55 yrs | 93 | 71.54 |
| Gender-male | 90 | 69.23 |
| Hypertension | 93 | 71.54 |
| Diabetes | 60 | 46.15 |
| Cigarette smoking | 20 | 15.38 |
| IHD | 15 | 11.54 |
| Other cardiac diseases | 11 | 8.46 |
| Alcohol | 12 | 9.23 |
| Anaemia | 6 | 4.62 |
| Atrial fibrillation | 6 | 4.62 |
| Dyslipidemia | 5 | 3.85 |
| **Number of medications received** |  |  |
| ≤5 | 9 | 6.92 |
| 6-10 | 22 | 16.92 |
| 11-15 | 30 | 23.08 |
| 16-20 | 30 | 23.08 |
| >20 | 39 | 30 |

| **Length of hospital stay(in days)** |  |  |
| --- | --- | --- |
| 0-5 | 26 | 20 |
| 6-10 | 43 | 33.08 |
| >10 | 61 | 46.92 |

### Type of Drug related problems identified

A total of 510 DRPs were identified from the sample population with an average incidence of 3.92 DRPs per patient. Prevalence of DRPs was found to be more in Hemorrhagic cases than in Ischemic cases and the prevalence of DRPs was found to be more in males than in females. Among all the DRPs identified drug drug interaction was found to be the highest accounting to 23.53% of total (**Table 2**).

**Table 2:**
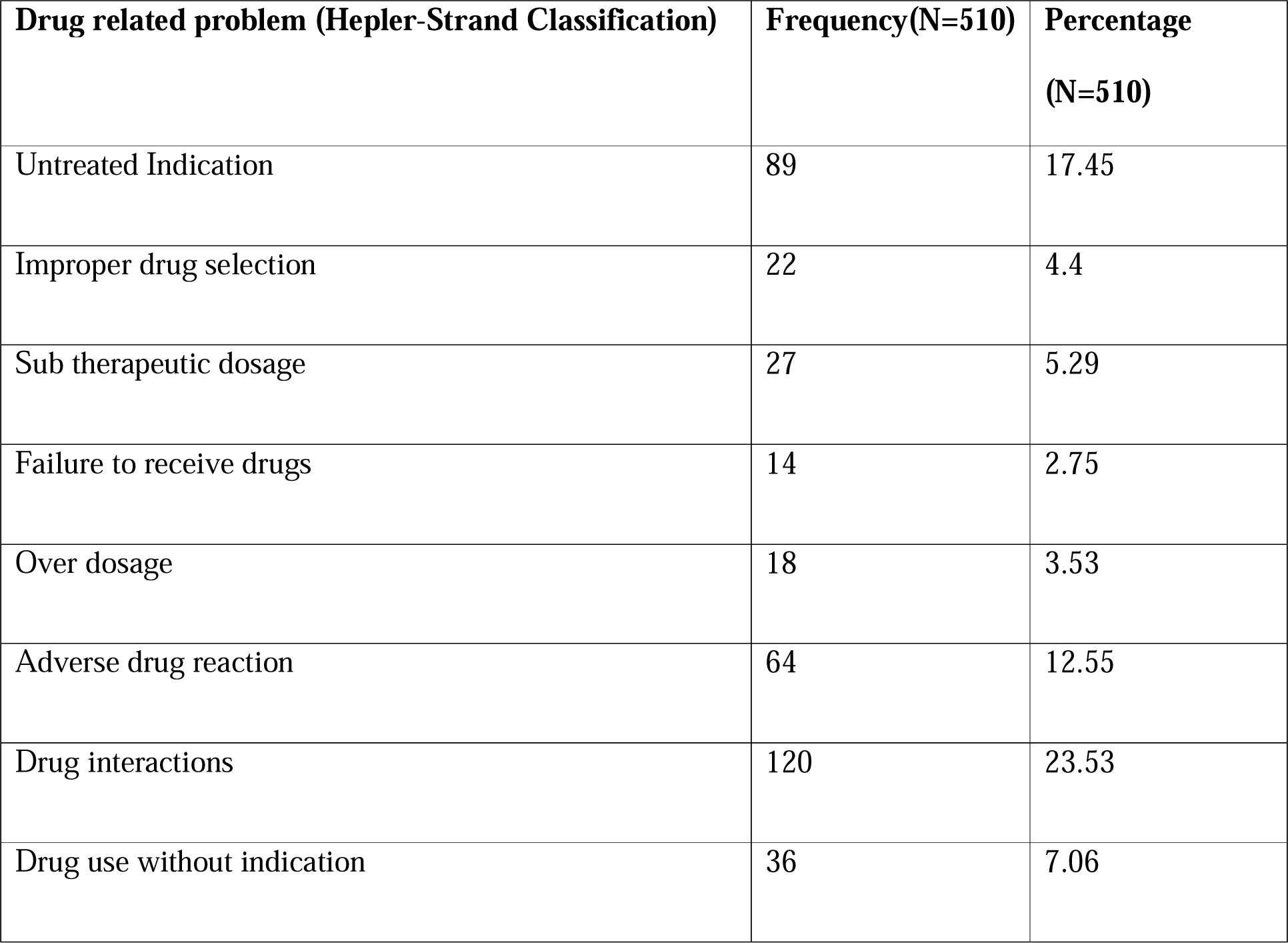
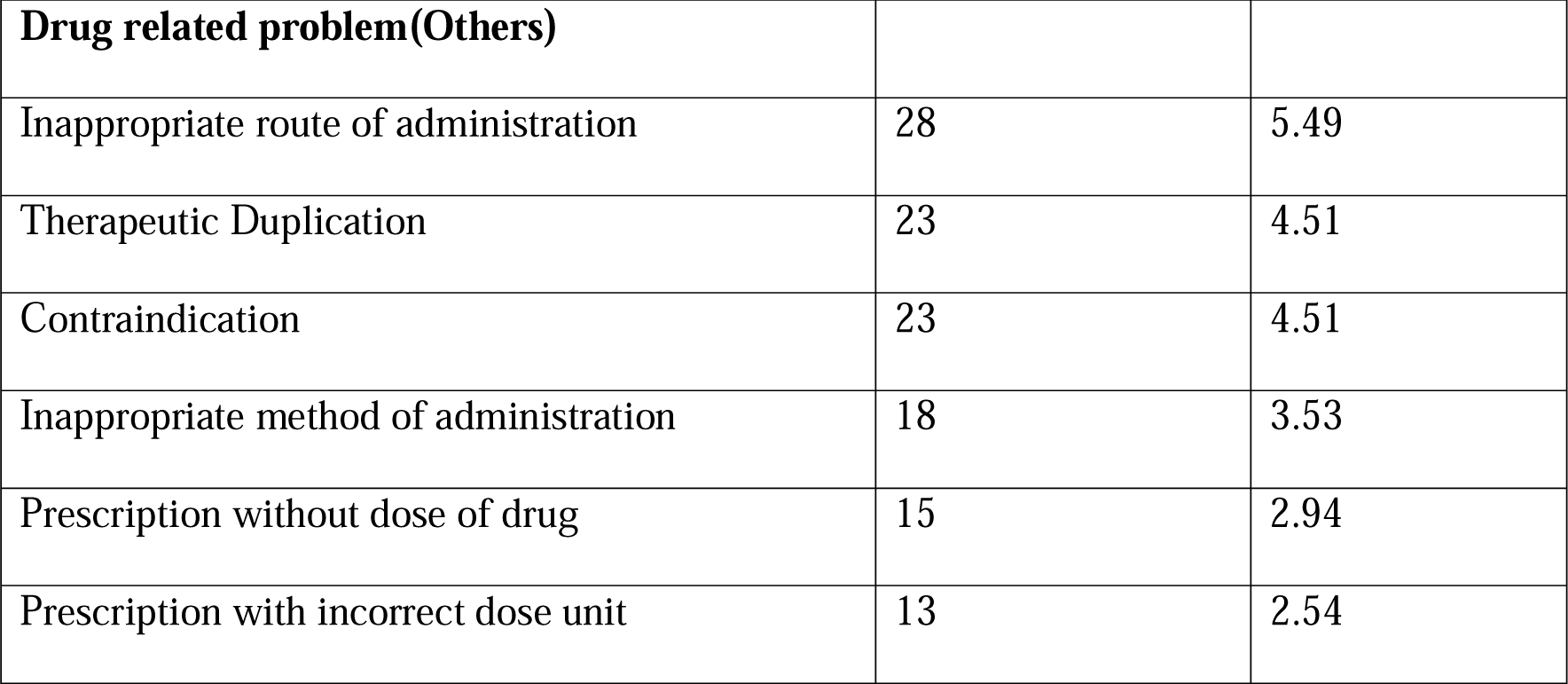
Type of Drug related problems identified.

### Drug-drug interactions

A total of 120 drug drug interactions due to 67 combinations of drugs occurred.80% of drug drug interactions were major,18.3 % moderate and 1.6% minor.The most common was the interaction between aspirin and clopidogrel which occurred 18 times. Excluding repeated drug interactions others are mentioned (**Table 3**)

**Table 3:**
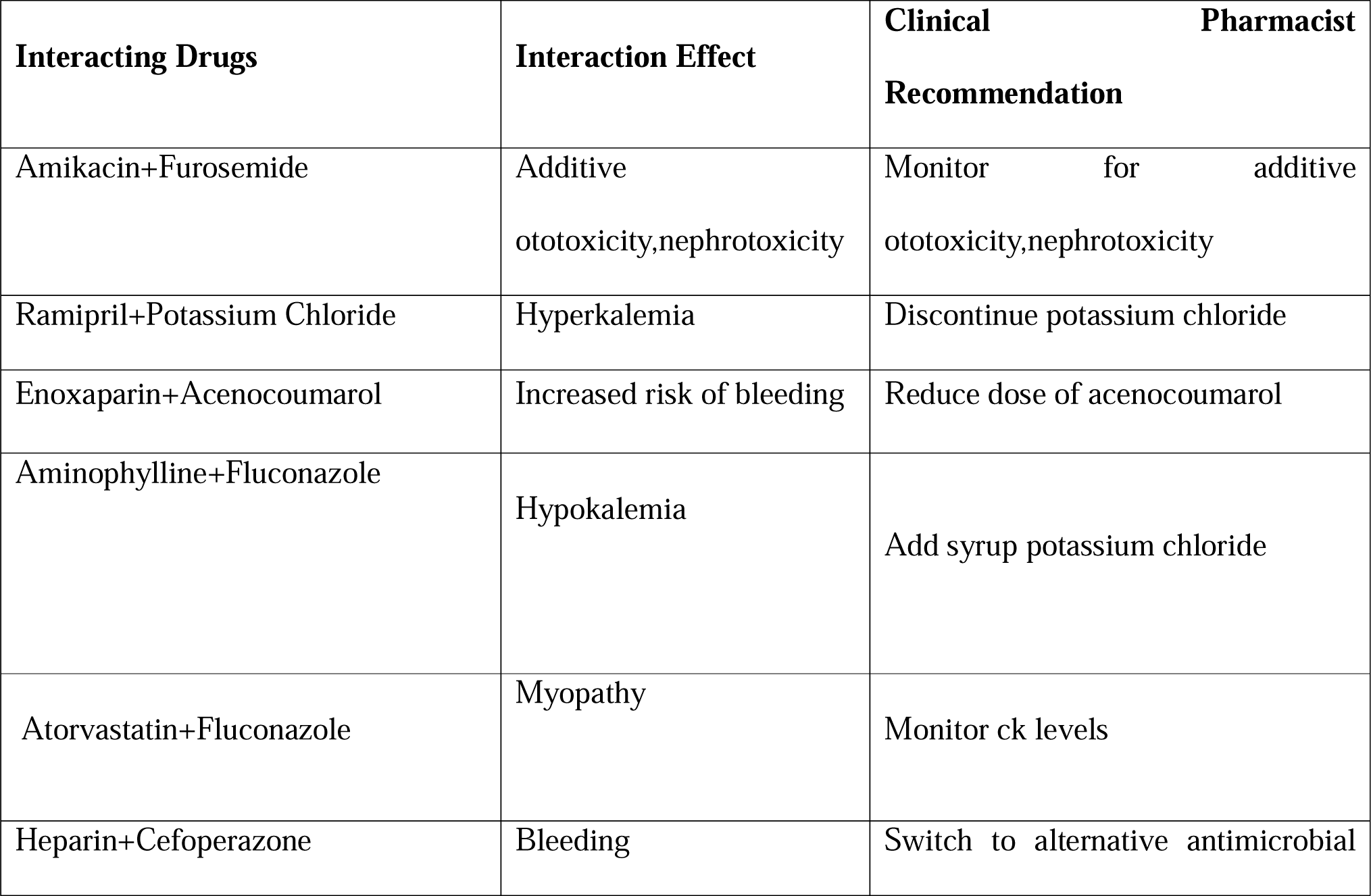
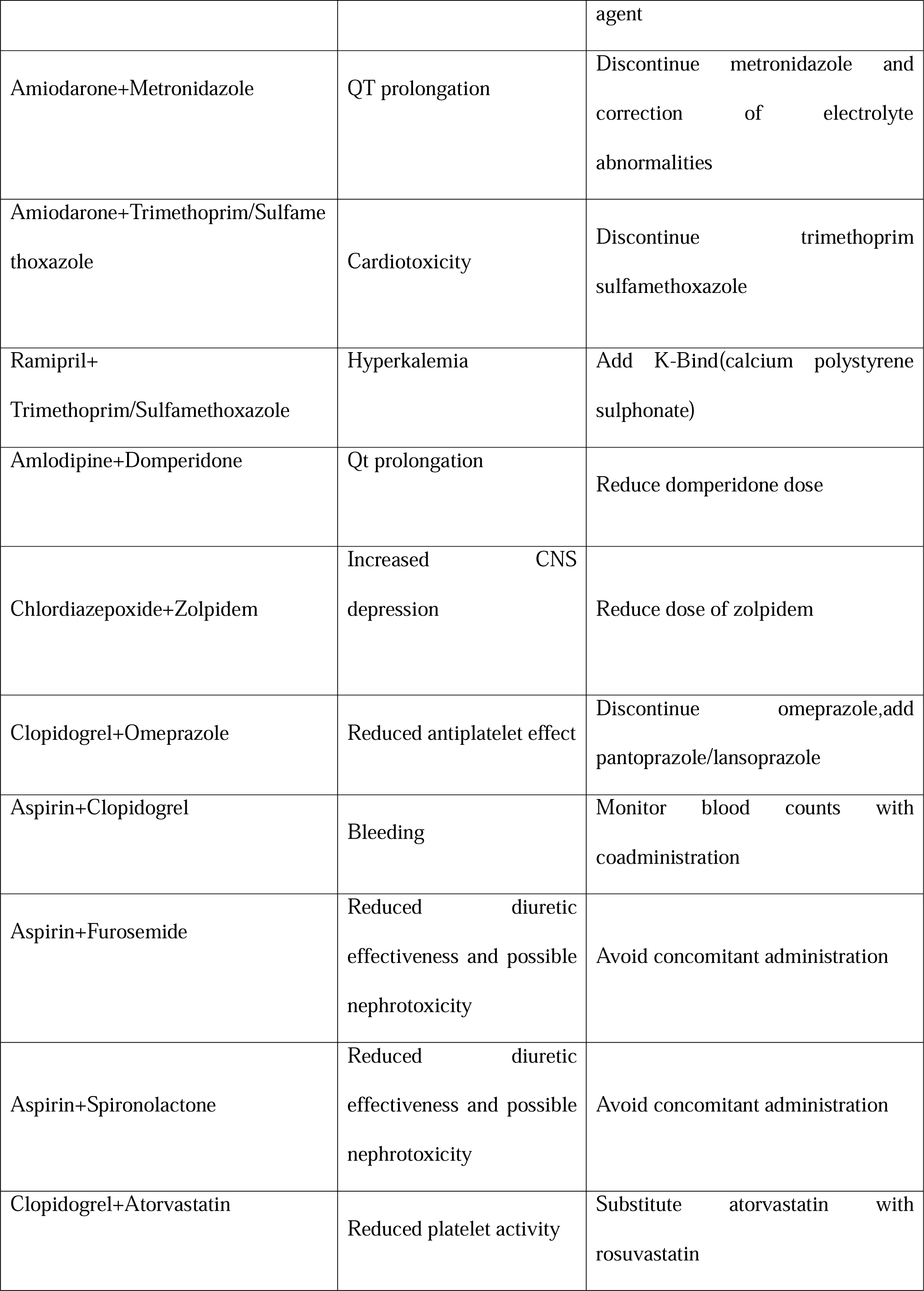
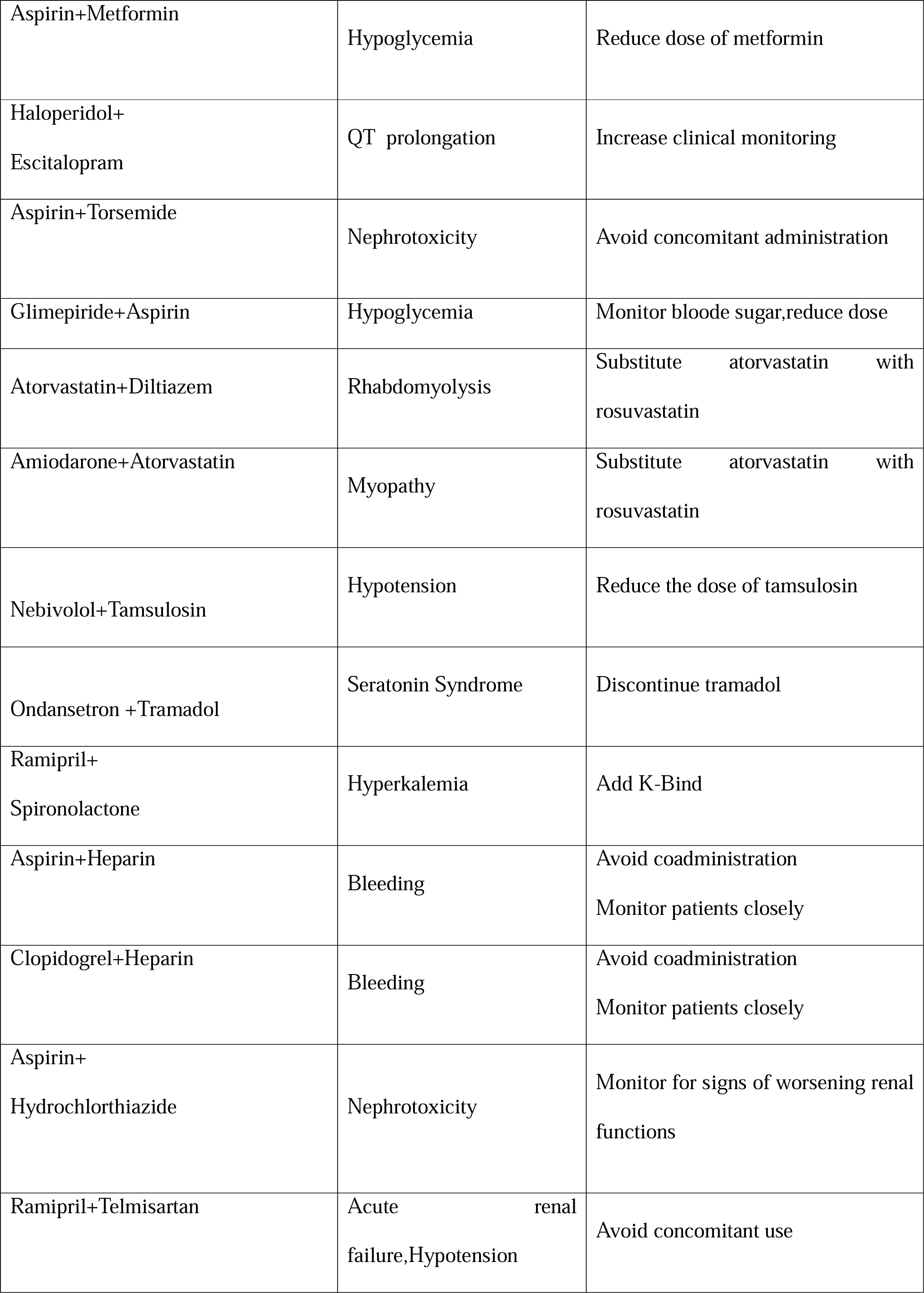
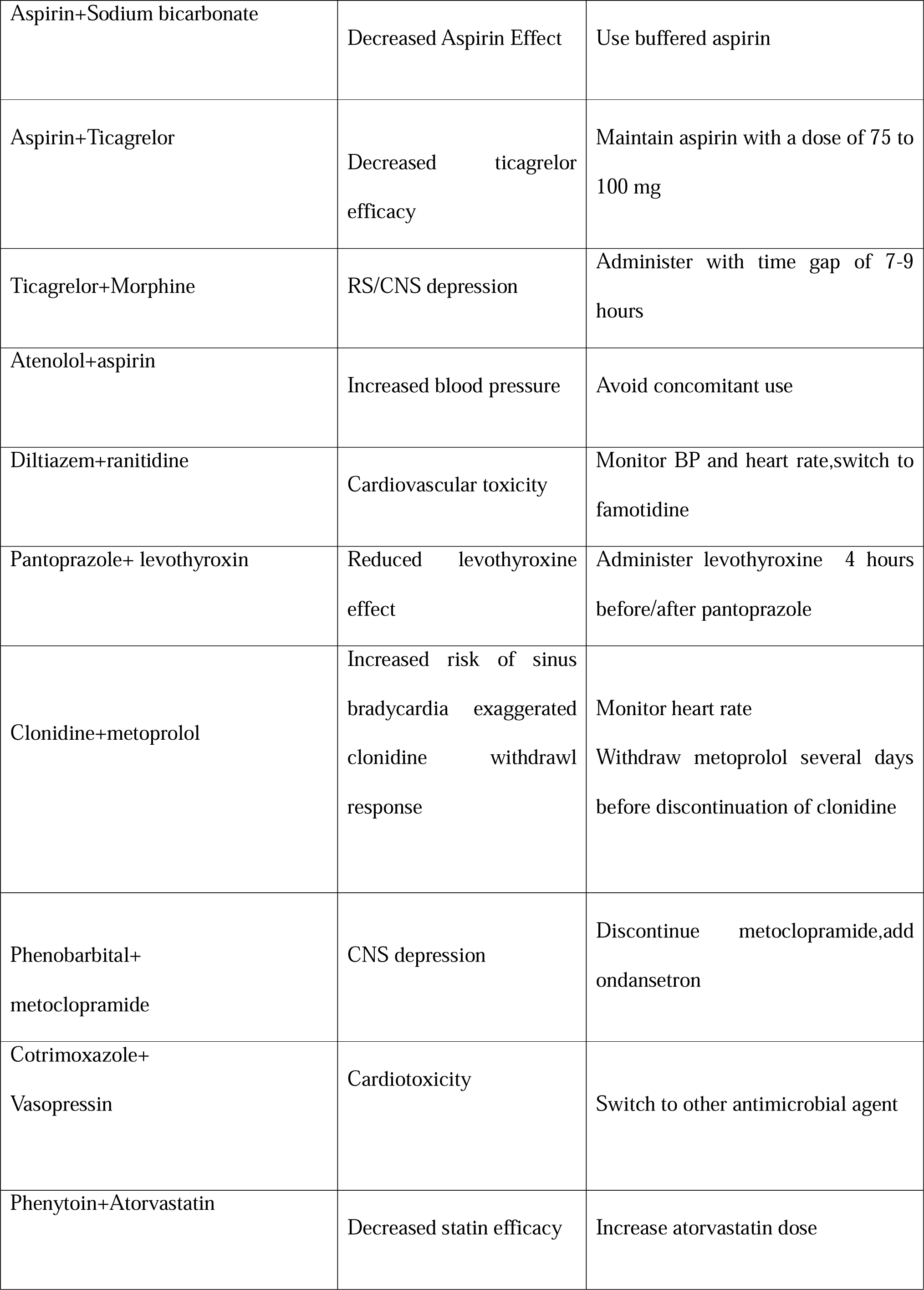
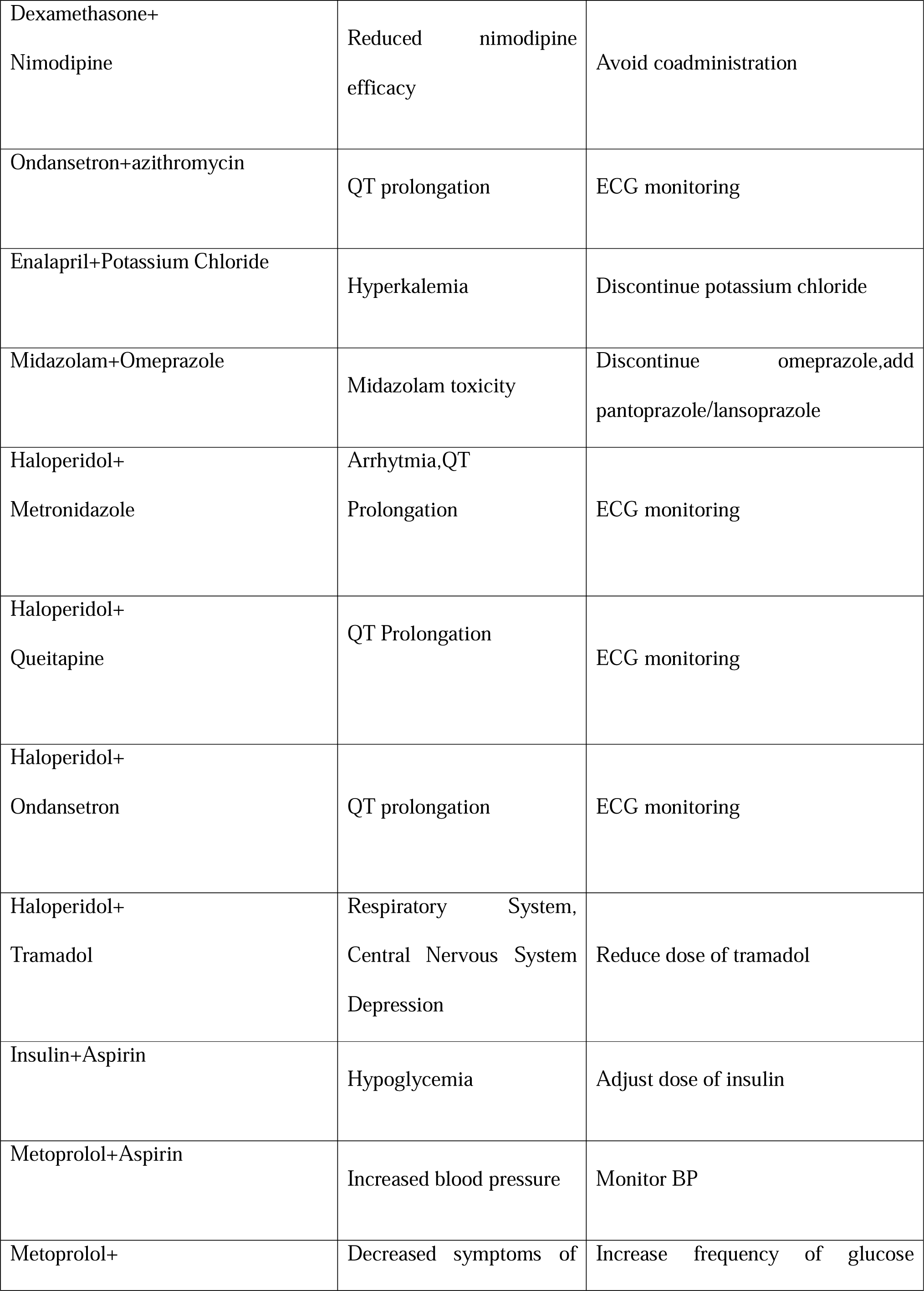
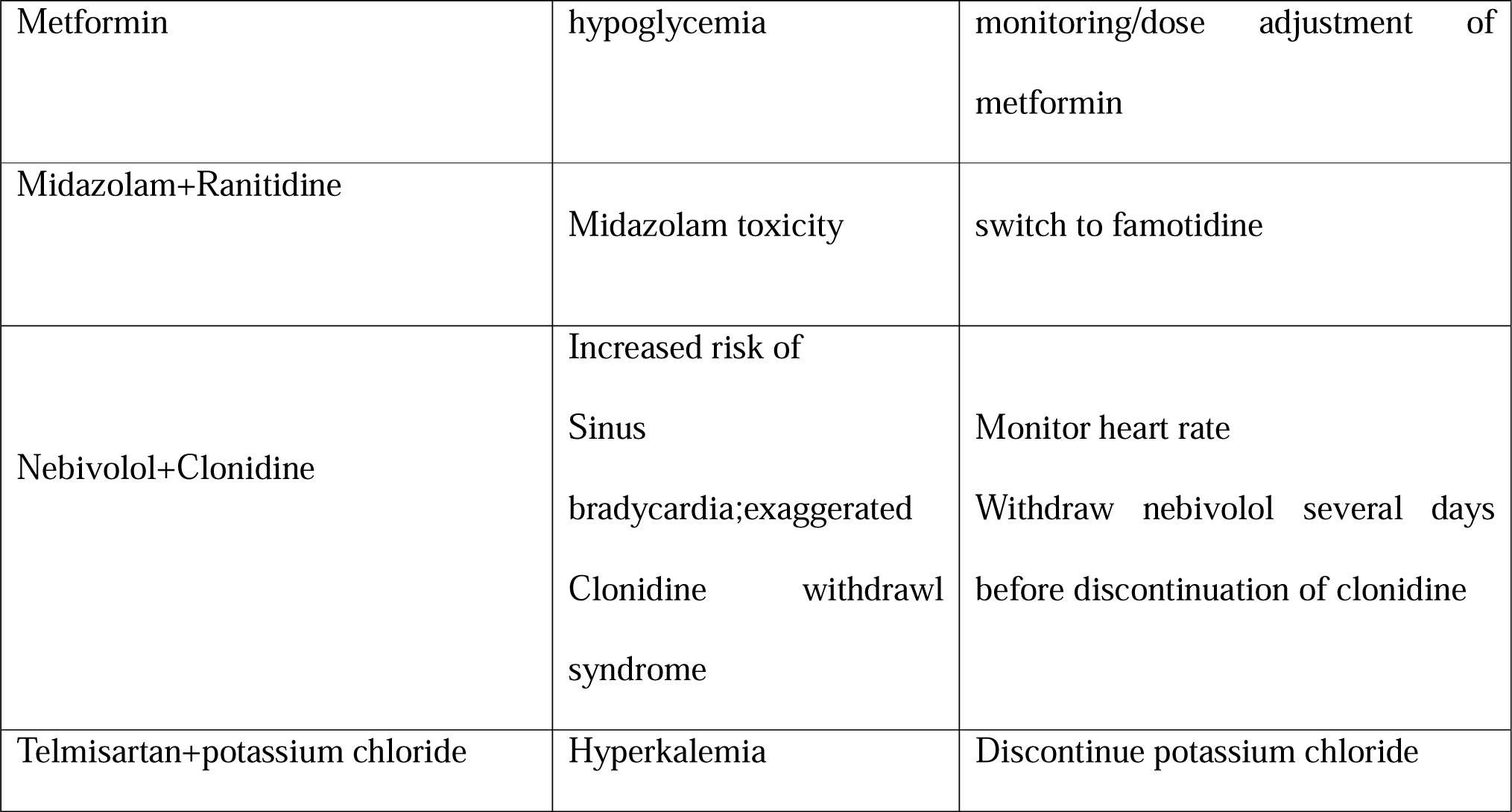
Drug Drug Interactions.

### Adverse drug reaction

A total of 64 ADRs caused by 33 different kind of drugs was observed. All the identified ADRs were assessed according to WHO probability scale. 39.06% of identified ADRs belonged to the category ‘certain’. out of 33 drugs 14 drugs(42.42%) caused ‘certain’ ADRS. Majority (60.94 %) of ADR belonged to the category ‘probable’(**Table 4**).

**Table 4:** Certain ADRs observed in sample population.

| Drug | Adr | Number Of Reports |
| --- | --- | --- |
| Doxycycline | Acute kidney injury | 1 |
| Meropenem | Loose stools | 1 |
|  | Fever | 1 |
|  | Acute kidney injury | 2 |
| Colistin | Diarrhea | 2 |
| Clonidine | Depression | 1 |
| Aspirin | GI bleed | 1 |
| Ceftriaxone | Allergic hives | 1 |
| Fentanyl | Bradycardia | 1 |
| Haloperidol | Hypotension | 2 |
|  | Blurring of vision | 1 |
|  | Xerostomia | 2 |
| Quetiapine | Drowsiness | 1 |
|  | Increased liver enzymes | 1 |
| Heparin | Melena | 1 |
| Insulin | Hypoglycemia | 2 |
| Piperacillin<br>tazobactam | Acute kidney injury | 1 |
| Paracetamol | LFT elevation | 1 |
| Telmisartan | Hyperkalemia | 2 |

### Recommendations made

Suggestions were provided to consultants, resident doctors and nurses. A high acceptance of recommendations (97.36%) was found in the study where changes in therapy were made (68.13) after the clinical pharmacist’s recommendations. Out of 510 DRPs, suggestions were made for 455 DRPs (**Table 5**).

**Table 5:**
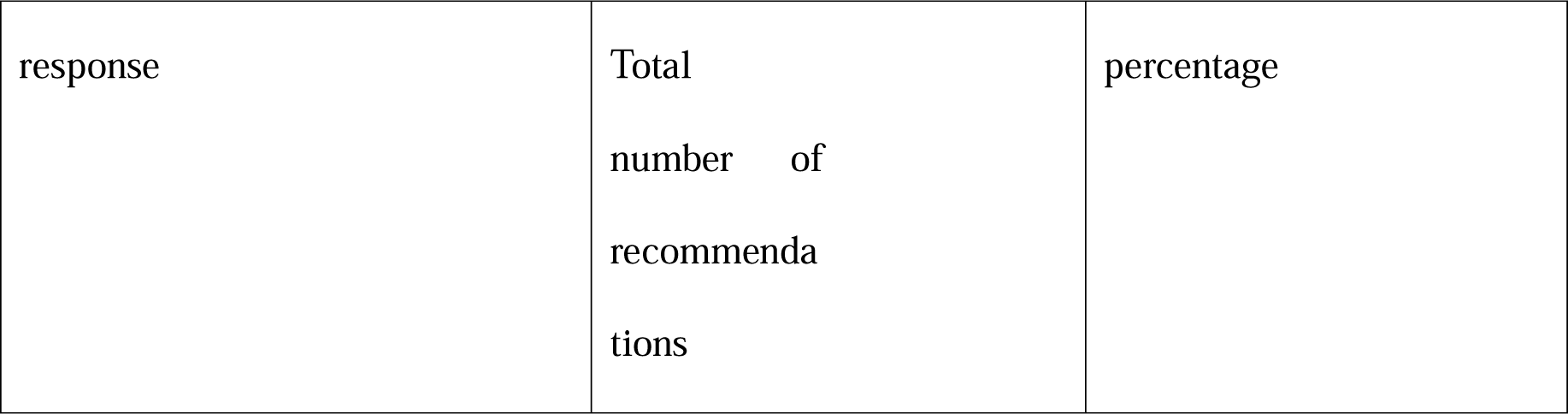
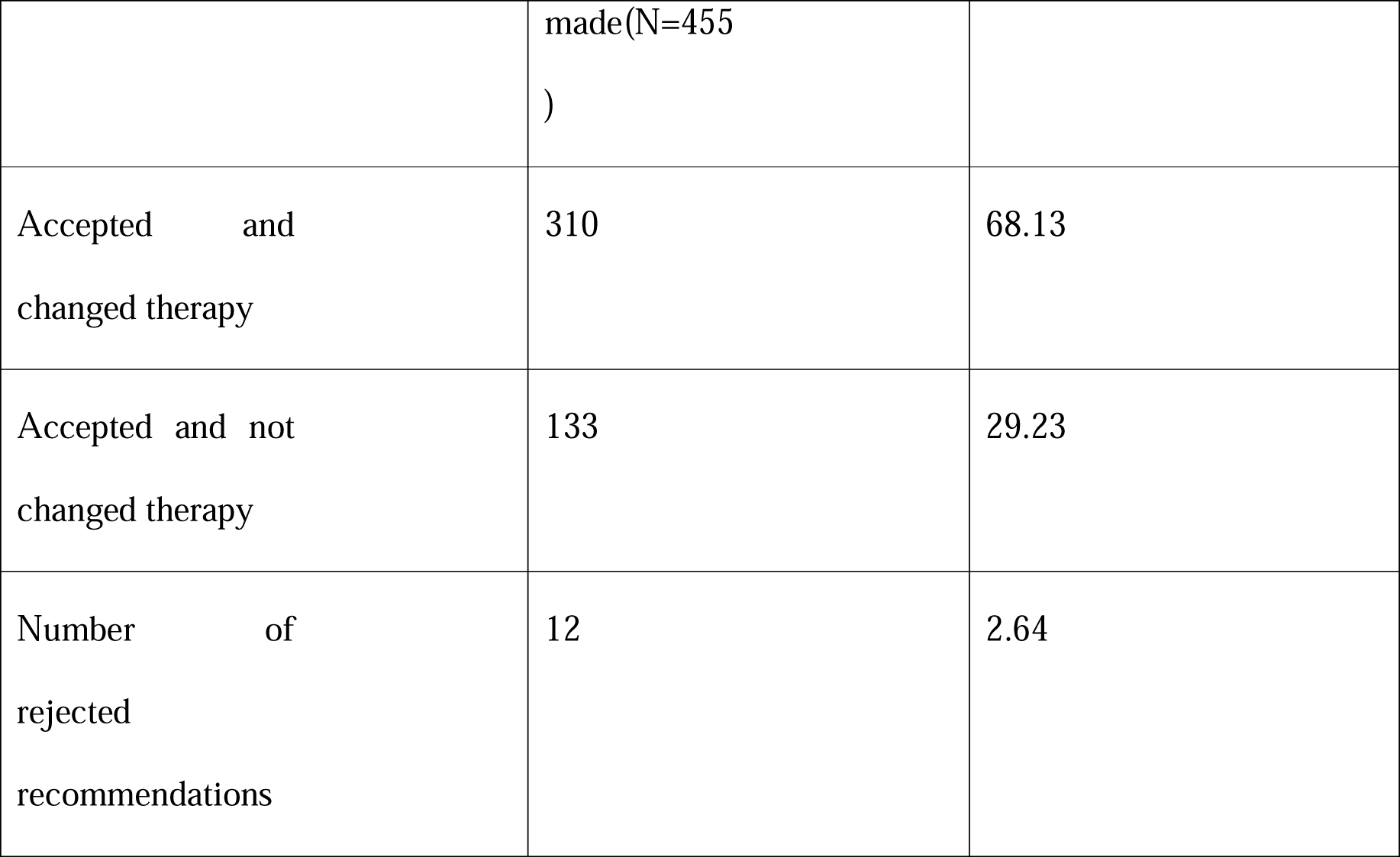
Clinical pharmacist’s recommendations and responses.

## DISCUSSION

A retrospective evaluation based on chart review performed in emergency department of a tertiary care centre revealed a 19% hospital incidence rate of medication errors.^14^According to a scientific statement by American Stroke Association (ASA) the mean length of hospital stay was 3 times longer in stroke patients with medical adverse events (including medication errors) than among patients who did not develop an adverse event^15^. One of the best trials to analyze and evaluate the DRPs involved in practice in hospital settings, and to test this specific DRP classification in the practice setting, is a prospective study.^16^ From the data gathered, the demographic information, clinical information, therapy administered, and types of drug-related concerns were examined. The admitted stroke patients received medicines like anti platelet agents, antihyperlipidemic agents, anticoagulants, antihypertensives, anti-diabetic drugs, anti-psychotic etc., The data showed that 39 individuals received more than 20 drugs, which was the most, overall. Among the stroke patients admitted, 70(53.85%), 44(33.85%), 16(12.3%) patients were admitted with ischemic stroke, hemorrhagic stroke and TIA respectively. Out of which, 90 (69.23%) were males and 30 (30.77) were females. Among the study population, majority (35.38%) of the patients were in the age group of 61-70 years. The demographic details of our study is concordant with the study results obtained by A T Celin et al, Divya Gopineni et al^9,16^. The study identified that hypertension is the major and frequent preventable risk factor followed by diabetes. These results were concordant with that of Vurumadla S et al.^17^ In our study majority of patients were diagnosed with ischaemic stroke followed by hemorrhagic which was in accordance with the study results of R P Eapen et al^18^.

A total of 510 DRPs were detected in 130 patients with an average incidence rate of 3.92 per patient. This was found to be higher than similar studies with comparable sample size^11^. DRPs were frequently seen in patients receiving more than 6 drugs. A 2002 national survey concluded that 50% of the population who took 5 or more medications developed DRPs^18^. The study conducted by Vinks TH et,al found that DRPs frequently occurred in adults over 65 years of age using 6 or more drugs concomitantly^19^. Similar results were obtained by the studies of A T celin ET al^9^. Predominant DRPs identified in our study was DDI followed by untreated indication and ADR. DDI contributed to 23.53 % of total DRPs. This observation was supported with the studies conducted by A T celin et al(25 %) and Yvonne Koh (34.8%)^9,19^. Untreated indications accounted for 17.45% of total DRPs. This was not consistent with the available published literatures. ADRs accounted for 12.55 % of total. Majority belonged to the category probable of the WHO probability scale proportion of ADR’s to the total DRP were comparable to the results by Celin.et.all^9^ in which all ADRs observed ADRs belonged to the category probable contrary to our study. Whereas a study conducted by Divya Gopineni et,al found 4 ADRs among the 264 DRPs and ADRs was not categorized as WHO probability scale.^11^ Drug use without indication was the fourth highest DRP. This was not in line with similar studies. Although published data’s are limited, the available information suggests that medication errors are common among patients hospitalized for acute stroke. With the obtained data our study tried to classify all the observed DRPs from prescribing to administration of the drug by Using Hepler-Strand Classification. Therefore, an elaborate picture of all possible DRPs were provided. The acceptance rate of recommendations was found to be very high (97.36%).In 68 % cases there was a change in drug therapy. These results were similar to the study by Celin et al where acceptance rate was 97 % and change in therapy was 70 %^9^.

Clinical Pharmacist involvement in patient-care rounds has been found to increase identification and tracking of DRPs and reduce the rate of occurrence of DRPs. Therefore, Clinical pharmacists can give proactive patient care through reactive recommendations and can also provide services to other healthcare professionals, such as drug information, to help them make effective therapeutic decisions.

## STRENGTHS AND LIMITATIONS

This study suggests that DRPs are extremely prevalent in a stroke unit. Some of these DRPs are distinctive due to stroke victims’ characteristics. Clinical pharmacists have the ability to effectively recognize and treat DRPs in a stroke unit. The high acceptance rate of the pharmacist’s assistance can be a sign of its high caliber in our study. However, this study has few limitations. The results of this study may not apply to other hospitals or neurological, General Medicine care units because it was carried out in a single academic tertiary care hospital. Clinical pharmacist’s recommendations has a long-term effects on patient outcomes were not assessed since they were outside the scope of this study. Future studies should concentrate on assessing the effects of clinical pharmacist’s impact over the long run for patients in a randomized controlled trial. The absence of control in our study is still another significant flaw, because of which the types and occurrences of DRPs in a stroke unit with and without clinical pharmacists could not be compared.

## CONCLUSION

The involvement of clinical pharmacists as a key member of the inpatient healthcare team is critical in minimizing medication errors in patients with acute stroke. Development of a standardized approach in prescribing the medications for common indications through protocols, critical pathways, and preprinted order forms or computerized order sets can minimize DRPs. This study helped to identify medication related discrepancies or issues that could be improved to help increase the quality of care for patients. Early detection and resolution of DRP may improve the therapeutic outcomes in stroke patients. This also helps in preventing complications and unnecessary hospitalization, high cost of treatment and deaths among stroke patients.

## Supporting information

Title page

## Data Availability

All data produced in the present study are available upon reasonable request to the authors

